# Mental Health of Medical Workers During the COVID-19 Pandemic in Russia: Results of a Cross-Sectional Study

**DOI:** 10.1101/2020.07.27.20162610

**Authors:** E.V. Bachilo, J.B. Barylnik, A.A. Shuldyakov, A.A. Efremov, D.E. Novikov

## Abstract

This is the first study in Russia regarding the mental health of medical workers during a pandemic. In this survey, the study of medical workers during pandemic COVID-19 in Russia reported high rates of symptoms of depression and anxiety. There is a higher risk of developing symptoms of anxiety and depression among young employees working directly in high-risk areas of the infection. Protecting medical workers is an important component of public health measures for addressing the COVID-19 pandemic. Special measures to improve the mental health of medical workers should be based on their needs. Special attention needs to be paid to young medical workers working in areas with a high risk of infection.

**Highlights:**

- medical workers are at high risk of developing mental disorders during a pandemic COVID-19;
- the overall prevalence of GAD (5 or more points on the GTD-7 scale) and depressive symptoms (5 or more points on the PHQ-9 scale) among medical workers in Russia during the pandemic period was 48.77% and 57.63% respectively;
- there is a higher risk of developing symptoms of anxiety and depression among young employees working directly in high-risk areas of the infection;
- the provision of personal protective equipment (69.2%), the optimization of the work and rest regime during the shift (58%), the support of management (52.7%) and relatives (51.1%) are the main areas that reduce the psycho-emotional stress of medical workers during a pandemic.

## Introduction

On January 30, 2020, the World Health Organization (WHO) announced that the emergence of a new coronavirus (2019-nCoV) was a public health emergency of international concern. On March 11, 2020, WHO announced the coronavirus pandemic [24]. To date, more than 3.5 million infected people worldwide have been identified. This is not the first outbreak of an infectious disease in the world. However, apparently, it will become the largest. Such outbreaks pose a rather serious public health problem and can affect not only the physical condition, but also the mental health of a person [6,12,16,26].

Previous studies have shown that during outbreaks of infectious diseases there is a wide spread of various negative psychological reactions, as well as the occurrence of symptoms of mental disorders. People may fear falling ill or dying, feel helpless and stigmatized [6]. Medical workers are also at risk of developing mental health problems [3,25]. Most often, medical workers expressed concerns about infecting themselves and their loved ones, colleagues, as well as a sense of self-doubt, stigmatization [1,17,23], noted high levels of stress, anxiety and symptoms of depression [14]. PTSD, depression and substance abuse can be long-term consequences of mental health problems experienced by medical workers during outbreaks of an infectious disease [4,11].

Obviously, medical workers are highly stressed and at high risk of adverse mental health consequences during the outbreak of COVID-19 [21]. The reasons for this are the following: a long working day, the risk of infection, lack of protective equipment, loneliness, physical fatigue and separation from families [7]. A number of studies have now been published that show the spectrum of mental health problems among medical workers during the COVID-19 pandemic [8,13,15,20]. Mostly, studies are presented by Chinese authors. In the majority of works, the main problems noted are: symptoms of anxiety disorder, depression, PTSD, sleep disturbances and excessive stress [8,13,15,20]. In Russia, such studies have not been conducted. This is the first study evaluating the mental health of Russian medical workers during the COVID-19 pandemic.

So, medical workers contacting with patients with COVID-19 obviously need certain interventions to maintain mental health. These interventions should be based on their needs [2].

The aim of the work was to assess the mental health of medical workers during the pandemic, to identify potential risk factors and needs in the field of psychosocial support. We hope that the results of our study will help to formulate recommendations that are effective and appropriate for the current situation for medical workers in conditions of increased psycho-emotional stress during the outbreak of COVID-19.

## 2. Materials and methods

### 2.1 Study Design and Participants

A survey has been compiled for health professionals and conducted via the Internet.

The survey was conducted among medical workers and non-medical health workers. A link to the survey was distributed in specialized groups on social networks, as well as via e-mail to the medical community.

The questionnaire was tested on 15 respondents who gave feedback regarding the understanding of each question in the questionnaire, after which the questions were adjusted for a clearer understanding of the respondents.

All respondents were able to familiarize themselves with informed consent prior to the start of the survey and could finish answering questions at any time.

### 2.2 Data collection

Participants answered questions anonymously, on the Internet from April 21, 2020 to May 18, 2020. Each participant filled out the demographic and social parts of the survey (including gender, age, children, education, specialty, marital status), standardized questionnaires questionnaire generalized anxiety disorder - GAD-7, the Depression Scale of the Human Health Assessment Questionnaire PHQ-9, part of questions about self-perceived health status, as well as a block of questions devoted to the most optimal ways to reduce the psychological impact of a pandemic on the mental health of medical workers.

In total, the survey involved 812 respondents from 77 regions of Russia.

#### 2.2.1. Demographic and social data

Basic demographic data include occupation (doctor, nurse, technical nurses, non-medical workers who work in the healthcare system), gender (male or female), age (20-25; 26-29; 30-39; 40-49; 50-59; 60 and older), education (incomplete secondary, secondary, college degree, incomplete higher, BA, MA, higher (specialty), PhD, MD); marital status (single / unmarried, in a civil marriage, unmarried, divorced, widow / widower, in a relationship); whether there are children (yes, no); specialty.

#### 2.2.2. Mental health assessment

We used 2 scales (Russian version of the scales) to assess the mental health status of medical workers. The 9-item Patient Health Questionnaire (PHQ-9) and the 7-item Generalized Anxiety Disorder (GAD-7) were used to evaluate symptoms of depression and anxiety. The PHQ-9 is a self-report measure used to assess the severity of depression, with the total scores categorized as follows: no depression (0), minimal (1–4), mild depression (5–9), moderate depression (10–14), severe depression (15–19) or extremely severe depression (20-27) [9,10]. The GAD-7 is a self-rated scale to evaluate the severity of anxiety and has good reliability and validity. The total scores are categorized as follows: minimal/no anxiety (0–4), mild anxiety (5– 9), moderate anxiety (10–14), or severe anxiety (15–21) [22].

#### 2.2.3. Self-perceived health status

Health status was determined by asking participants to compare their current health status to their health status before the outbreak of COVID-19: How do you perceive your current health status compared to your health status before the outbreak? (answer options included: getting better, almost unchanged, worse, or much worse).

We also asked the respondents to evaluate their quality of sleep (from 0 to 10, where 0 - sleep is grossly violated, 10 - quality of sleep has not changed).

#### 2.2.4. Psychological support needs for health professionals

One of the objectives of the survey was to obtain information from medical professionals regarding their needs for psychological assistance. The following questions were suggested:

- Do you think that healthcare providers assisting patients with COVID-19 need psychological support? (Yes or no);
- What psychological support options do you consider most appropriate for healthcare workers providing care for patients with COVID-19? (Answer options: Provision of personal protective equipment, optimization of the work /rest regime during the work shift, management’s support, support for loved ones, support for colleagues, the ability to isolate themselves from their loved ones in order to prevent infection, organizing the assistance of specialists (psychologists, psychotherapists, psychiatrists), doing exercise to stabilize their psycho-emotional state, access to hotline for psychological support. Also we gave the opportunity to offer a different answer).
- Which of the following, in modern conditions, seems to you the most optimal for preserving your psychological state among medical workers? (Answer options: Brochures / booklets with tips on how to relieve stress, reduce anxiety; Psychological support telephone hotline; Direct consultation of a specialist (psychologist, psychiatrist, psychotherapist); Short-term group lessons with a psychologist before a shift; Sites and apps to relieve stress and reduce anxiety. Also we gave the opportunity to offer a different answer).

According to the results of the survey, similar answers were grouped together.

### 2.3 Ethical aspects

The study was conducted in accordance with the Helsinki Declaration and was approved by the ethics committee of the Scientific and Educational Center for Psychotherapy and Clinical Psychology. The participants could familiarize themselves with the informed consent prior to the start of the survey. The participants could stop doing the survey at any time without explanation.

### 2.3. Statistical analysis

A descriptive analysis of the data was carried out to understand the demographic and social characteristics of the interviewed medical workers in Russia and their needs for psychological support during the COVID-19 pandemic. To compare differences in the prevalence of symptoms of anxiety and depression between groups the chi-square test (χ2) was used. P values of less than 0.01 were considered statistically significant.

Internal consistency using the PHQ-9 and GAD-7 questionnaires was evaluated using Cronbach alpha statistics. Descriptive statistics for quantitative variables are presented as mean values (standard deviation) and medians (1st and 3rd quartiles). Descriptive statistics for categorical variables are presented as the number of cases per category (percentage).

To compare quantitative variables, the Kruskel-Wallis test was used. For qualitative ones, the χ2 test with approximation of the distribution of statistics using the Monte Carlo method was used. The differences were considered statistically significant at p <0.05. The association of potential risk factors with the outcomes studied (anxiety and depression levels) was evaluated using one- and multi-factor proportional odds models. The estimates obtained from the questionnaires were not categorized; the models used quantitative-dependent variables [5]. The association of risk factors with the outcomes was considered statistically significant at p <0.05.

Analysis and visualization of the obtained data were carried out using the statistical computing environment R 3.6.3 (R Foundation for Statistical Computing, Vienna, Austria), the Statistical Package for Social Sciences (SPSS) version 24.0, Microsoft Excel.

## 3. Results

### 3.1. Demographic and social data

812 respondents participated in the survey. Of the 812 responding participants, 641 (79%) were doctors, 138 (17%) were nurses, 7 (0,9%) were technical nurses and 25 (3,1%) were non-medical workers. Most participants were women (658 [81%]), aged 20 to 49 years (644 [79,4%]), married (470 [57,9%]), with educational level of higher specialist (545 [67,1%]) and 334 (41,1%) workers were directly involved in assisting patients with COVID-19 (Table 1).

**Table 1.** Demographic and social characteristics of respondents (N = 812).

|  | n (%) |
| --- | --- |
|  | 812 (100) |
| **Gender** |  |
| Male | 154 (19) |
| Female | 658 (81) |
| **Age** |  |
| 20-25 | 106 (13,1) |
| 26-29 | 119 (14,7) |
| 30-39 | 226 (27,8) |
| 40-49 | 193 (23,8) |
| 50-59 | 123 (15,1) |
| 60 и более | 45 (5,5) |
| **Education** |  |
| incomplete secondary | 1 (0,1) |
| secondary | 7 (0,9) |
| college degree | 123 (15,1) |
| incomplete higher | 32 (3,9) |
| BA | 16 (2) |
| MA | 14 (1,7) |
| higher specialist | 545 (67,1) |
| PhD | 64 (7,9) |
| MD | 10 (1,2) |
| **Marital status** |  |
| single, not married | 157 (19,3) |
| in a civil marriage | 54 (6,7) |
| married | 470 (57,9) |
| divorced | 73 (9) |
| widow / widower | 20 (2,5) |
| in a relationship | 38 (4,7) |
| **Children** |  |
| Yes | 524 (64,5) |
| No | 288 (35,5) |
| **Occupation** |  |
| Doctor | 641 (79) |
| Nurse | 138 (17) |
| Technical nurses | 7 (0,9) |
| Non-medical worker | 25 (3,1) |
| **Work in high-risk areas** |  |
| Yes | 334 (41,1) |
| No | 478 (58,9) |

### 3.2. Mental health assessment

The prevalence of GAD, depressive symptoms stratified by gender, age, and occupation were shown in Table 2 and Table 3.

**Table 2.** Prevalence of GAD and depressive symptoms during COVID-19 outbreak in Russia’s medical workers stratified by gender and age (N = 812).

| | Total<br>(N=812)<br>n(%) | Male<br>(N=154)<br>n(%) | Female<br>(N=658)<br>n(%) | $\chi^2$ | p-value | 20-25<br>(N=106)<br>n(%) | 26-29<br>(N=119)<br>n(%) | 30-39<br>(N=226)<br>n(%) | 40-49<br>(N=193)<br>n(%) | 50-59<br>(N=123)<br>n(%) | 60 and older<br>(N=45)<br>n(%) | $\chi^2$ | p-value |
| --- | --- | --- | --- | --- | --- | --- | --- | --- | --- | --- | --- | --- | --- |
| **GAD-7** |  |  |  | **3,14** | **0,37** |  |  |  |  |  |  | **19,72** | **0,00019** |
| minimal/<br>no anxiety | 416<br>(51,23%) | 86<br>(55,84%) | 329<br>(50%) |  |  | 48<br>(45,28%) | 51<br>(42,85%) | 114<br>(50,44%) | 100<br>(51,81%) | 72<br>(58,54%) | 31<br>(68,88%) |  |  |
| mild anxiety | 263<br>(32,39%) | 46<br>(29,87%) | 214<br>(32,52%) |  |  | 37<br>(34,91%) | 45(37,82%) | 71<br>(31,41%) | 67<br>(34,72%) | 30<br>(24,39%) | 13(28,89%) |  |  |
| moderate anxiety | 87<br>(10,71%) | 17<br>(11,04%) | 70<br>(10,63%) |  |  | 14<br>(13,21%) | 15<br>(12,6%) | 26<br>(11,5%) | 16<br>(8,29%) | 15<br>(12,20%) | 1<br>(2,22%) |  |  |
| severe anxiety<br>**PHQ-9** | 46<br>(5,67%) | 5<br>(3,25%) | 41<br>(6,23%) | **5,19** | **0,39** | 7<br>(6,60%) | 8<br>(6,72%) | 15<br>(6,64%) | 10<br>(5,18%) | 6<br>(4,88%) | 0<br>(0%) | **47,22** | **<0.001** |
| no depression | 101<br>(12,43%) | 16<br>(10,39%) | 85<br>(12,92%) |  |  | 8<br>(7,55%) | 10<br>(8,4%) | 24<br>(10,62%) | 27<br>(13,99%) | 20<br>(16,26%) | 12<br>(26,67%) |  |  |
| minimal depression | 243<br>(29,93%) | 53<br>(34,42%) | 190<br>(28,88%) |  |  | 21<br>(19,81%) | 33<br>(27,73%) | 62<br>(27,43%) | 66<br>(34,19%) | 43<br>(34,96%) | 18<br>(40%) |  |  |
| mild depression | 220<br>(27,09%) | 41<br>(26,62%) | 179<br>(27,2%) |  |  | 32<br>(30,19%) | 31<br>(26,05%) | 65<br>(28,76%) | 49<br>(25,39%) | 32<br>(26,02%) | 11<br>(24,44%) |  |  |
| moderate depression | 141<br>(17,36%) | 29<br>(18,82%) | 112<br>(17,02%) |  |  | 23<br>(27,7%) | 29<br>(24,37%) | 44<br>(19,47%) | 28<br>(14,51%) | 13<br>(10,57%) | 4<br>(8,89%) |  |  |
| severe depression | 78<br>(9,61%) | 13<br>(8,44%) | 65<br>(9,88%) |  |  | 17<br>(16,04%) | 12<br>(10,08%) | 21<br>(9,29%) | 15<br>(7,77%) | 13<br>(10,57%) | 0<br>(0%) |  |  |
| extremely severe depression | 29<br>(3,57%) | 2<br>(1,3%) | 27<br>(4,1%) |  |  | 5<br>(4,72%) | 4<br>(3,36%) | 10<br>(4,42%) | 8<br>(4,15%) | 2<br>(1,63%) | 0<br>(0%) |  |  |

Abbreviations: GAD-7, the 7-item Generalized Anxiety Disorder; PHQ-9, The 9-item Patient Health Questionnaire.

**Table 3.**
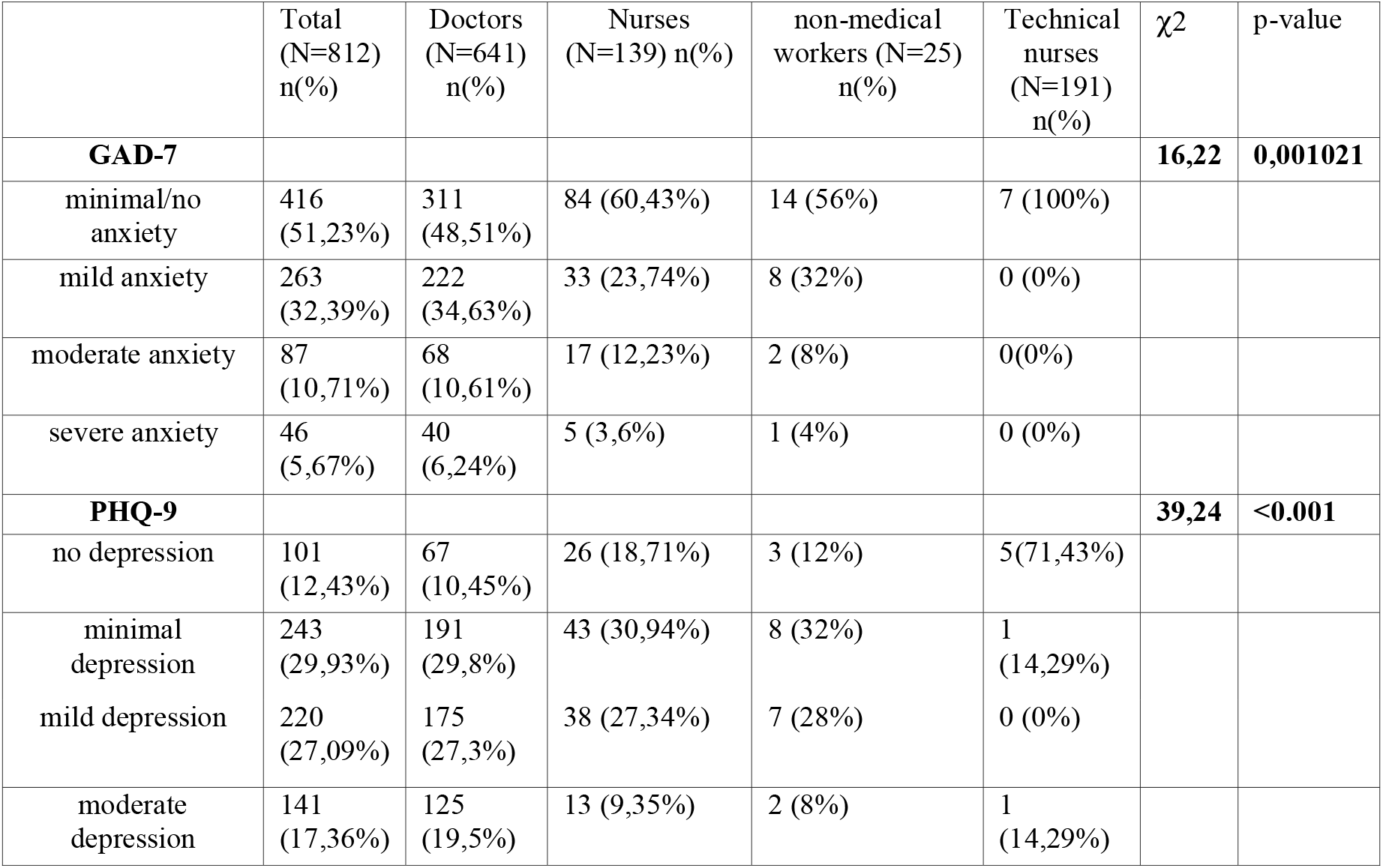
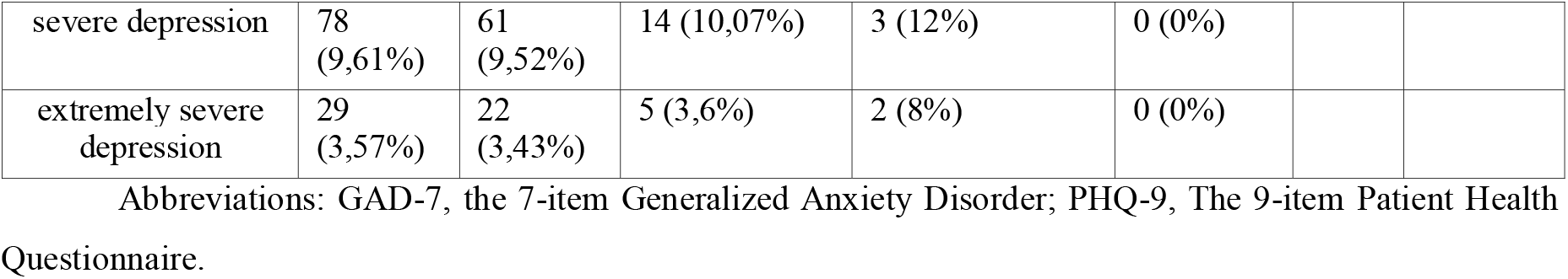
Prevalence of GAD and depressive symptoms during COVID-19 outbreak in Russia’s medical workers stratified by occupation (N = 812).

| | Total<br>(N=812)<br>n(%) | Doctors<br>(N=641)<br>n(%) | Nurses<br>(N=139) n(%) | non-medical<br>workers (N=25)<br>n(%) | Technical<br>nurses<br>(N=191)<br>n(%) | $\chi^2$ | p-value |
| --- | --- | --- | --- | --- | --- | --- | --- |
| **GAD-7** |  |  |  |  |  | **16,22** | **0,001021** |
| minimal/no anxiety | 416<br>(51,23%) | 311<br>(48,51%) | 84 (60,43%) | 14 (56%) | 7 (100%) |  |  |
| mild anxiety | 263<br>(32,39%) | 222<br>(34,63%) | 33 (23,74%) | 8 (32%) | 0 (0%) |  |  |
| moderate anxiety | 87<br>(10,71%) | 68<br>(10,61%) | 17 (12,23%) | 2 (8%) | 0(0%) |  |  |
| severe anxiety | 46<br>(5,67%) | 40<br>(6,24%) | 5 (3,6%) | 1 (4%) | 0 (0%) |  |  |
| **PHQ-9** |  |  |  |  |  | **39,24** | **<0.001** |
| no depression | 101<br>(12,43%) | 67<br>(10,45%) | 26 (18,71%) | 3 (12%) | 5(71,43%) |  |  |
| minimal depression | 243<br>(29,93%) | 191<br>(29,8%) | 43 (30,94%) | 8 (32%) | 1<br>(14,29%) |  |  |
| mild depression | 220<br>(27,09%) | 175<br>(27,3%) | 38 (27,34%) | 7 (28%) | 0 (0%) |  |  |
| moderate depression | 141<br>(17,36%) | 125<br>(19,5%) | 13 (9,35%) | 2 (8%) | 1<br>(14,29%) |  |  |
| severe depression | 78<br>(9,61%) | 61<br>(9,52%) | 14 (10,07%) | 3 (12%) | 0 (0%) |  |  |
| extremely severe depression | 29<br>(3,57%) | 22<br>(3,43%) | 5 (3,6%) | 2 (8%) | 0 (0%) |  |  |

Abbreviations: GAD-7, the 7-item Generalized Anxiety Disorder; PHQ-9, The 9-item Patient Health Questionnaire.

The overall prevalence of GAD (5 or more points on the GAD-7 scale) and depressive symptoms (5 or more points on the PHQ-9 scale) were 48.77% and 57.63% respectively. There was no statistically significant difference in the prevalence of GAD and depressive symptoms by gender (P > 0,01, as shown in Table 2). The analysis of the prevalence of symptoms of anxiety and depression by age revealed statistically significant differences (P<0.01). Higher rates of medium and high levels of anxiety were characteristic of younger people (in the range of 20–39 years old), while respondents over 50 years of age in most cases showed no or minimal level of anxiety. The vast majority of the respondents from the group with mild symptoms of depression belonged to the age range of 20 - 39 years old; among respondents with severe symptoms were participants from the age groups of 20-25, 26-29, 50-59 years old (Table 2). There was no significant difference in the prevalence of anxiety symptoms by occupation (P> 0.01, as shown in Table 3). However, there was a difference in the prevalence of symptoms of depression depending on the occupation (P<0.01). Thus, the overall prevalence of depressive symptoms (5 or more points on the PHQ-9 scale) among doctors was 59.75%, among nurses - 50.36%, among non-medical workers - 56%, among technical nurses - 14.29%.

### 3.3. Self-perceived health status

Table 4 shows a subjective assessment of one’s health status compared to the period before the pandemic. The vast majority of the respondents, 78.1%, indicated that there were no changes. 15.9% reported deterioration. Table 5 presents the results of a subjective assessment of sleep quality. 32.4% (261 participants) noted that almost no sleep disturbance occured. 37.4% respondents (305 people) indicated sleep disturbances from 0 to 5 points.

**Table 4.** Subjective assessment of health status compared to the period before the outbreak of COVID-19 (N = 812).

|  | n (%) |
| --- | --- |
| almost unchanged | 634 (78,1) |
| worse | 129 (15,9) |
| getting better | 26 (3,2) |
| much worse | 23 (2,8) |

**Table 5.**
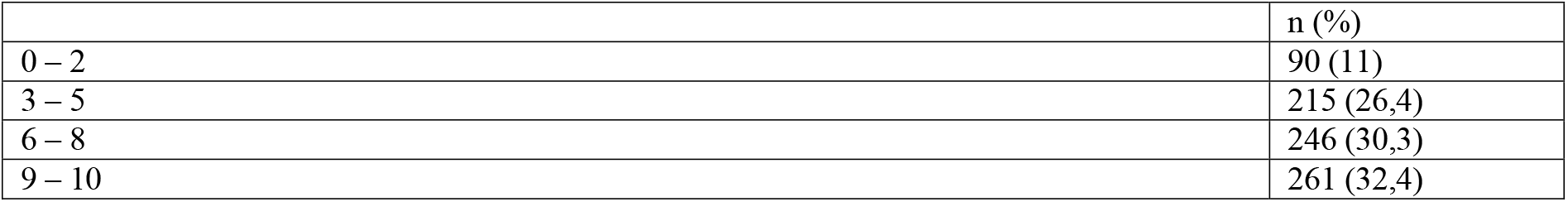
Subjective assessment of sleep quality (N = 812).

### 3.4. Association of influence factors with anxiety and depressive symptoms in medical workers during COVID-19 outbreak

The associations of potential influence factors with GAD and depressive symptoms in medical workers during COVID-19 outbreak were presented in Table 6.

**Table 6.** Estimates of the size of the effect (with corresponding 95% confidence intervals) obtained in one- and multi-factor proportional odds models (N = 812).

|  |  | GAD-7 |  | PHQ-9 |  |
| --- | --- | --- | --- | --- | --- |
|  |  | pOR (CI) | pOR<sub>adj</sub> (CI) | pOR (CI) | pOR<sub>adj</sub> (CI) |
| Age | 20-25 | 1,24 (0,83–1,86) | 1,13 (0,67–1,88) | 1,56 (1,04–2,34) | 1,33 (0,79–2,23) |
|  | 26-29 | 1,29 (0,87–1,92) | 1,15 (0,75–1,75) | 1,16 (0,79–1,71) | 1,02 (0,67–1,55) |
|  | 30-39 | 1,00 | 1,00 | 1,00 | 1,00 |
|  | 40-49 | 0,91 (0,65–1,27) | 0,96 (0,68–1,36) | 0,70 (0,50–0,99) | 0,71 (0,50–1,01) |
|  | 50-59 | 0,75 (0,51–1,11) | 0,78 (0,52–1,16) | 0,59 (0,40–0,87) | 0,62 (0,42–0,93) |
|  | 60 and more | 0,33 (0,19–0,59) | 0,37 (0,21–0,68) | 0,26 (0,15–0,45) | 0,31 (0,17–0,54) |
| Gender | F | 1,00 | 1,00 | 1,00 | 1,00 |
|  | M | 1,01 (0,75–1,37) | 0,87 (0,63–1,19) | 0,96 (0,71–1,29) | 0,79 (0,58–1,09) |
|  | married | **1,00** | 1,00 | **1,00** | 1,00 |
|  | divorced | **1,27 (0,83–1,95)** | 1,17 (0,75–1,80) | **1,37 (0,89–2,11)** | 1,34 (0,87–2,07) |
|  | single, not married | **1,28 (0,93–1,76)** | 0,96 (0,64–1,43) | **1,81 (1,32–2,49)** | 1,36 (0,91–2,04) |
|  | widow / widower | **0,24 (0,11–0,54)** | 0,41 (0,17–0,97) | **0,17 (0,08–0,37)** | 0,30 (0,12–0,71) |
|  | in relationship | **1,20 (0,69–2,08)** | 0,85 (0,46–1,56) | **1,39 (0,78–2,48)** | 1,04 (0,57–1,92) |
|  | in a civil marriage | **1,20 (0,72–2,00)** | 1,14 (0,67–1,96) | **1,41 (0,84–2,35)** | 1,32 (0,77–2,26) |
| Children | Yes | **1,00** | 1,00 | **1,00** | 1,00 |
|  | No | **1,48 (1,15–1,90)** | 1,18 (0,81–1,71) | **1,83 (1,42–2,36)** | 1,06 (0,72–1,57) |
| Education | incomplete secondary | **0,20 (0,01–3,25)** | 0,16 (0,01–2,78) | **1,42 (0,09–22,91)** | 0,77 (0,05–12,94) |
|  | secondary | **0,23 (0,06–0,93)** | 0,32 (0,06–1,61) | **0,21 (0,04–1,08)** | 0,30 (0,05–1,82) |
|  | college degree | **0,48 (0,34–0,68)** | 0,55 (0,27–1,12) | **0,49 (0,35–0,70)** | 0,48 (0,24–0,99) |
|  | incomplete higher | **1,55 (0,85–2,83)** | 1,42 (0,63–3,20) | **1,67 (0,88–3,17)** | 1,03 (0,44–2,41) |
|  | BA | **0,48 (0,20–1,16)** | 0,40 (0,16–0,98) | **0,83 (0,34–2,02)** | 0,70 (0,28–1,72) |
|  | higher specialist | **1,00** | 1,00 | **1,00** | 1,00 |
|  | MA | **1,08 (0,40–2,93)** | 1,03 (0,40–2,68) | **1,11 (0,40–3,08)** | 1,06 (0,40–2,75) |
|  | PhD | **0,81 (0,51–1,29)** | 0,91 (0,57–1,47) | **0,71 (0,45–1,10)** | 0,91 (0,58–1,44) |
|  | MD | **0,68 (0,24–1,93)** | 1,06 (0,36–3,11) | **0,78 (0,26–2,31)** | 1,28 (0,42–3,90) |
| Occupation | Non-medical worker | **1,00 (0,50–1,99)** | 1,12 (0,54–2,29) | **1,11 (0,54–2,29)** | 1,16 (0,54–2,48) |
|  | Technical nurse | **0,12 (0,03–0,49)** | 0,26 (0,05–1,30) | **0,05 (0,01–0,27)** | 0,14 (0,02–0,90) |
|  | Nurse | **0,62 (0,45–0,87)** | 0,89 (0,44–1,81) | **0,64 (0,46–0,89)** | 0,99 (0,49–2,01) |
|  | Doctor | **1,00** | 1,00 | **1,00** | 1,00 |
| Work in high risk areas | No | **1,00** | **1,00** | **1,00** | **1,00** |
|  | Yes | **1,48 (1,16–1,90)** | **1,43 (1,10–1,84)** | **1,50 (1,18–1,92)** | **1,41 (1,09–1,82)** |

Abbreviations: GAD-7, the 7-item Generalized Anxiety Disorder; PHQ-9, The 9-item Patient Health Questionnaire; *OR*, odds ratio; *ORadj*, adjusted odds ratio; 95% *CI*, 95% confidence interval.

When using the PHQ-9 questionnaire, Cronbach’s alpha was 0.875 (95% CI: 0.861– 0.887), while using GAD-7 it was 0.902 (95% CI: 0.889–0.913), thus we can conclude that the obtained results are reliable.

In the univariate models, age, marital status, having children, education, occupation and work in high-risk areas were associated with anxiety levels and the severity of symptoms of depression. So, higher levels of anxiety were characteristic of: young people (20 - 29 years old) (20-25 years: *OR*=1.24, 95% *CI*: 0.83-1.86; 26-29 years: *OR*=1.29, 95% *CI*: 0.87-1.92), who do not have children (*OR*=1.48, 95% *CI*: 1.15-1.90), have incomplete higher education (students) (*OR*=1.55, 95% *CI*: 0.85-2.83), working in areas with a high risk of infection (*OR*=1.48, 95% *CI*: 1.16-1.90). Doctors and non-medical workers had approximately the same associations between the work performed and the level of anxiety. More pronounced symptoms of depression were associated with young age (20 - 29 years) (20-25 years: *OR*=1.56, 95% *CI*: 1.04-2.34; 26-29 years: *OR*=1.16, 95% *CI*: 0.79-1.71), absence of children (*OR*=1.83, 95% *CI*: 1.42-2.36), education (incomplete secondary (*OR*=1.42, 95% *CI*: 0.09-22.91), incomplete higher (*OR*=1.67, 95% *CI*: 0.88-3.17), MA (*OR*=1.11, 95% *CI*: 0.40-3.08), performance in the health care system of work not related to the direct provision of medical care (non-medical workers) (*OR*=1.11, 95% *CI*: 0.54-2.29), work in the zone of increased risk of infection (*OR*=1.5, 95% *CI*: 1.18-1.92).

In the multivariate models, the above associations weakened but there was still statistical difference. In proportional odds models, it was found that old age (over 60) is statistically associated with lower levels of symptoms of anxiety *(OR*=0.37, 95% *CI*: 0.21-0.68) and depression (*OR*=0.31, 95% *CI*: 0.17-0.54). Gender was not associated with the outcomes studied. Work in high-risk areas of the infection is statistically significantly associated with high levels of symptoms of anxiety (*OR*=1.43, 95% *CI*: 1.10-1.84) and depression (*OR*=1.41, 95% *CI*: 1.09-1.82). Age and work in areas with an increased risk of infection have been independent predictors of anxiety and depressive disorders.

So, on the basis of multifactorial models of proportional chances, we can conclude that there is a higher risk of developing symptoms of anxiety and depression among young employees working directly in high-risk areas of the infection.

### 3.5. Psychological support needs for health professionals

Table 7 and Table 8 provide data on the most optimal directions and options for supporting healthcare providers during a pandemic. In general, 87.4% of respondents said that psychological support is necessary for medical workers. The provision of personal protective equipment (69.2%), the optimization of the work and rest regime during the shift (58%), the support of management (52.7%) and relatives (51.1%) - these are the main areas (for more than 50 percent of respondents) that reduce the psycho-emotional stress of medical workers during a pandemic. The most popular options for support were direct consultation with a specialist (psychologist, psychiatrist, psychotherapist) (38.8%), short group sessions before a shift with a psychologist (35.5%), sites and applications that help relieve stress and reduce anxiety (10.2%), brochures/booklets with tips on how to relieve stress and reduce anxiety (4.7%), call a telephone hotline for psychological support (3.3%), organization of work and rest (1.8%).

**Table 7.** The most optimal directions of assistance to medical workers to maintain their psychological state during a pandemic (N = 812).

|  | n (%) |
| --- | --- |
| **Answers to choose from** |  |
| Personal protective equipment | 562 (69,2) |
| Optimization of the regime of work and rest during the work shift | 471 (58) |
| Leadership support | 428 (52,7) |
| Support from loved ones | 464 (51,1) |
| Peer support | 329 (40,5) |
| The ability to isolate yourself from your loved ones in order to prevent infection | 310 (38,2) |
| Organization of specialist assistance (psychologists, psychotherapists, psychiatrists) | 175 (21,6) |
| Performing exercises to stabilize your psycho-emotional state | 154 (19) |
| The ability to contact the telephone line for psychological support | 44 (5,4) |
| **Other ideas of respondents** |  |
| Material incentive | 15 (1,85) |
| Compensation in case of illness | 1 (0,1) |
| Legal support | 1 (0,12) |
| Decent attitude of society and patients | 3 (0,37) |
| Appeal to religion | 2 (0,25) |
| Removal of restrictive measures in connection with COVID-19 | 2 (0,25) |
| Clear algorithms of work | 1 (0,12) |

**Table 8.** The most optimal psychological support options for medical workers providing care to patients with COVID-19 according to the medical workers themselves (N = 812).

|  | n (%) |
| --- | --- |
| Direct consultation of a specialist (psychologist, psychiatrist, psychotherapist) | 315 (38,8) |
| Short-term group lessons before a shift with a psychologist | 287 (35,3) |
| Sites and apps to relieve stress and reduce anxiety | 83 (10,2) |
| Brochures/booklets with tips on how to relieve stress and reduce anxiety | 38 (4,7) |
| Psychological support telephone hotline | 27 (3,3) |
| Organization of work and rest | 15 (1,8) |
| Financial incentive | 6 (0,7) |
| Security | 4 (0,5) |
| Coronavirus information filtering | 4 (0,5) |
| I do not know | 3 (0,37) |
| Safety, Organization of work and rest | 3 (0,37) |
| Support from loved ones | 3 (0,37) |
| Support and communication with colleagues | 3 (0,37) |
| No answer | 2 (0,25) |
| Short-term individual lessons before the work shift | 1 (0,12) |
| Group lessons after the shift | 1 (0,12) |
| Organization of places for psychological relief | 1 (0,12) |
| Community support | 1 (0,12) |
| Spa treatment at the expense of the employer | 1 (0,12) |
| Physical and respiratory practices, fasting days | 1 (0,12) |
| Appeal to the Church and Religion | 1 (0,12) |
| Other (reading, learning languages) | 1 (0,12) |

## Discussion

This is the first study in Russia regarding the mental health of healthcare providers during a pandemic. 812 respondents participated in the survey. Of the 812 responding participants, 641 (79%) were doctors, 138 (17%) were nurses, 7 (0,9%) were technical nurses and 25 (3,1%) were non-medical workers. Most participants were women (658 [81%]), aged 20 to 49 years old (644 [79,4%]), married (470 [57,9%]), had an educational level of higher specialist (545 [67,1%]) and 334 (41,1%) workers were directly involved in assisting patients with COVID-19. The overall prevalence of GAD (5 or more points on the GTD-7 scale) and depressive symptoms (5 or more points on the PHQ-9 scale) was 48.77% and 57.63% respectively. Higher rates of medium and high levels of anxiety were characteristic of younger people (in the range of 20–39 years old), while respondents over 50 years of age in most cases showed no or minimal level of anxiety. The vast majority of respondents from the group with mild symptoms of depression belonged to the age range of 20 - 39 years old; among respondents with severe symptoms were participants from the age groups of 20-25, 26-29, 50-59 years old. 37.4% of participants reported poor sleep quality. Age and work in areas with an increased risk of infection have been independent predictors of anxiety and depressive disorders. There is a higher risk of developing symptoms of anxiety and depression among young employees working directly in high-risk areas of the infection. The provision of personal protective equipment (69.2%), the optimization of the work and rest regime during the shift (58%), the support of management (52.7%) and relatives (51.1%) are the main areas that reduce the psycho-emotional stress of medical workers during a pandemic. The most popular options for support were direct consultation with a specialist (psychologist, psychiatrist, psychotherapist) (38.8%), short group sessions before a shift with a psychologist (35.5%), sites and applications that help relieve stress and reduce anxiety (10.2%), brochures/booklets with tips on how to relieve stress and reduce anxiety (4.7%), call a telephone hotline for psychological support (3.3%), organization of work and rest (1.8%).

The results of this study are consistent with other studies of the mental health of medical workers during a pandemic in China and Italy. In our study, there was no significant difference in the prevalence of symptoms of anxiety and depression by gender, like in the study by Yun Chen et al. (2020) [27], while several other researchers have revealed such dependence [13,20]. Higher levels of anxiety and depression were characteristic of young professionals working in areas of high risk of infection. Young age and high levels of anxiety and depression are indicated in other studies of the mental health of medical workers [20].

Obviously, medical workers need support for their mental health during periods of pandemics, when psycho-emotional stress increases significantly. The needs of healthcare providers must be considered in order to provide the most effective support. It is necessary to elaborate in detail the model of rendering, directions and algorithms of this assistance.

One of the works illustrates the situation when a gap arises between the planned mental health services in a particular hospital and the actual needs of medical workers [2]. As a result, assistance directions were adjusted in accordance with requests. So, in our study, we have identified the basic needs and forms of support that will be most optimal for workers in the Russian healthcare system.

In China, a national guide to emergency psychological intervention during the outbreak of COVID-19 was developed, in which a special role is given to assisting medical workers [4,18]. In Russia, the Ministry of Healthcare sent an official letter to the regions with recommendations on the organization of psychological and psychotherapeutic care in connection with the spread of the new coronavirus infection COVID-19 to the population of the Russian Federation, including medical workers (letter No. 28-3 / I / 2-6111 of 05/07/2020). The Union of Specialists for Mental Health has developed recommendations for medical workers who are under increased psycho-emotional stress during the COVID-19 pandemic [19].

At the same time, this area of assistance to healthcare system specialists remains insufficiently developed due to the lack of research in this area, in particular among Russian medical workers. Thus, this study, conducted among medical workers from different regions, is the first such study in the Russian Federation and shows that a rather large proportion of medical workers (especially young adults) involved in the COVID-19 pandemic have mental health problems. The directions and forms of support indicated by the respondents themselves are presented, which can help in the formation of an aid system now and in the future, in the event of similar situations.

## Conclusion

### Limitations

This study has several limitations. First, since the data and relevant analyses presented here were derived from a cross-sectional design, it is difficult to make causal inferences. Secondly, the study was limited to COVID-19 outbreak, and we used a web-based survey method to avoid contracting the infection during an offline mode. Hence, causing the sampling of our study was voluntary and conducted by online. Therefore, the possibility of selection bias should be considered. Thirdly, due to the sudden occurrence of the disaster, we were unable to assess an individual’s psychological condition before the outbreak.

## Data Availability

All data was available

## Disclosures and acknowledgments

The authors declare that they have no competing interests. The authors have no sources of funding or other financial disclosures concerning the above article.

## Notes

### Competing Interest Statement

The authors have declared no competing interest.

### Author Declarations

All relevant ethical guidelines have been followed; any necessary IRB and / or ethics committee approvals have been obtained and details of the IRB / oversight body are included in the manuscript.

## References

1. Bai Y, Lin CC, Lin CY, Chen JY, Chue CM, Chou P. Survey of stress reactions among health care workers involved with the SARS outbreak. Psychiatr Serv. 2004; 55 (9): 1055–1057. https://doi.org/10.1176/appi.ps.55.9.1055

2. Chen Q., Liang M., Li Y., Guo J. et al.. Mental health care for medical staff in China during the COVID-19 outbreak. Lancet Psychiatry. 2020 Apr;7(4):e15–e16. https://doi.org/10.1016/S2215-0366(20)30078-X

3. Chong M.Y. Psychological impact of severe acute respiratory syndrome on health workers in a tertiary hospital. Br. J. Psychiat. J. Mental Sci. 2004;185:127–133;

4. Das N. Psychiatrist in post-COVID-19 era - Are we prepared? Asian J Psychiatr. 2020;51:102082. doi:10.1016/j.ajp.2020.102082

5. FE Harrell Jr. Regression modeling strategies: with applications to linear models, logistic regression, and survival analysis (2nd Edition). Springer. 2015. P 361–365

6. Hall R.C.W., Chapman M.J. The 1995 Kikwit Ebola outbreak: Lessons hospitals and physicians can apply to future viral epidemics. Gen. Hosp. Psychiatry. 2008;30:446–452. https://doi.org/10.1016/j.genhosppsych.2008.05.003

7. Kang L., Li Y., Hu S., Chen M., Yang C., Yang B.X., Wang Y., Hu J., Lai J., Ma X., Chen J., Guan L., Wang G., Ma H., Liu Z. The mental health of medical workers in Wuhan, China dealing with the 2019 novel coronavirus. Lancet Psychiatry. 2020 Mar;7(3):e14. https://doi.org/10.1016/S2215-0366(20)30047-X

8. Kang L, Ma S, Chen M, Yang J., Wang Y., Li R., Yao L., Bai H., Cai Z., Yang B.X., Hu S., Zhang K., Wang G., Ma C., Liu X. Impact on mental health and perceptions of psychological care among medical and nursing staff in Wuhan during the 2019 novel coronavirus disease outbreak: A cross-sectional study. Brain Behav Immun. 2020;S0889- 1591(20)30348-2. https://doi.org/10.1016/j.bbi.2020.03.028

9. Kocalevent R.D. Standardization of the depression screener patient health questionnaire (PHQ-9) in the general population. Gen. Hospital Psychiat. 2013; 35:551–555. https://doi.org/10.1016/j.genhosppsych.2013.04.006

10. Kroenke K, Spitzer RL. The PHQ-9: A new depression diagnostic and severity measure. Psychiatric Annals. 2002; 32(9):509–515;

11. Lancee WJ, Maunder RG, Goldbloom DS; Coauthors for the Impact of SARS Study. Prevalence of psychiatric disorders among Toronto hospital workers one to two years after the SARS outbreak. Psychiatr Serv. 2008;59(1):91–95. doi:10.1176/ps.2008.59.1.91;

12. Lau JT., Griffiths S., Choi KC., Tsui HY. Avoidance behaviors and negative psychological responses in the general population in the initial stage of the H1N1 pandemic in Hong Kong. BMC Infect Dis. 2010;10:139. https://doi.org/10.1186/1471-2334-10-139

13. Lai J, Ma S, Wang Y, et al. Factors Associated With Mental Health Outcomes Among Health Care Workers Exposed to Coronavirus Disease 2019. JAMA Netw Open. 2020;3(3):e203976. Published 2020 Mar 2. https://doi.org/10.1001/jamanetworkopen.2020.3976

14. Lee AM, Wong JG, McAlonan GM, et al.. Stress and psychological distress among SARS survivors 1 year after the outbreak. Can J Psychiatry. 2007; 52 (4): 233–240. https://doi.org/10.1177/070674370705200405

15. Liu S., Yang L., Zhang C., Xiang Y.T., Liu Z., Hu S., Zhang B. Online mental health services in China during the COVID-19 outbreak. Lancet Psychiatry. 2020;7(4):e17–e18. https://doi.org/10.1016/S2215-0366(20)30077-8

16. Mak W, Chu CM, Pan PC, Yiu MG, Ho SC, Chan VL. Risk factors for chronic post-traumatic stress disorder (PTSD) in SARS survivors. Gen Hosp Psychiat. 2010;32:590–598. https://doi.org/10.1016/j.genhosppsych.2010.07.007

17. Maunder R, Hunter J, Vincent L, et al.. The immediate psychological and occupational impact of the 2003 SARS outbreak in a teaching hospital. CMAJ. 2003; 168 (10): 1245–1251;

18. Rana W, Mukhtar S, Mukhtar S. Mental health of medical workers in Pakistan during the pandemic COVID-19 outbreak [published online ahead of print, 2020 Apr 7]. Asian J Psychiatr. 2020;51:102080. https://doi.org/10.1016/j.ajp.2020.102080;

19. Recommendations for medical workers who are in conditions of increased psycho-emotional stress during the COVID-19 pandemic. Accessed June 16, 2020. http://rosmededucation.ru/sites/default/files/files/pdf/Рекомендации%20для%20медраотников.pdf (In Russ);

20. Rodolfo Rossi, Valentina Socci et al. Mental health outcomes among front and second line health workers associated with the COVID-19 pandemic in Italy. MedRxiv. 2020.04.16.20067801. https://doi.org/10.1101/2020.04.16.20067801

21. Shigemura J., Ursano R.J., Morganstein J.C., Kurosawa M., Benedek D.M. Public responses to the novel 2019 coronavirus (2019 – nCoV): mental health consequences and target populations. Psychiatry Clin. Neurosci. 2020 Apr;74(4):281–282. https://doi.org/10.1111/pcn.12988

22. Spitzer R.L., Kroenke K., Janet B. W. Williams, Löwe B. A brief measure for assessing generalized anxiety disorder. Arch Intern Med. 2006;166(10):1092–1097. https://doi.org/10.1001/archinte.166.10.1092

23. Wong T.W., Yau J.K., Chan C. L. et al.. The psychological impact of severe acute respiratory syndrome outbreak on healthcare workers in emergency departments and how they cope. Eur J Emerg Med. 2005 Feb; 12 (1): 13–8. https://doi.org/10.1097/00063110-200502000-00005

24. World Health Organization. WHO Director-General’s opening remarks at the media briefing on COVID-19 - March 11, 2020;

25. Wu P. The psychological impact of the SARS epidemic on hospital employees in China: exposure, risk perception, and altruistic acceptance of risk. Canadian journal of psychiatry. Revue canadienne de psychiatrie. 2009;54:302–311. https://doi.org/10.1177/070674370905400504

26. Xiang YT, Yu X, Ungvari GS, Correl CU, Chiu HF. Outcomes of SARS survivors in China: not only physical and psychiatric co-morbidities. East Asian Arch Psychiatry. 2014;24:37–38. PMID: 24676486;

27. Yun Chen, Hao Zhou, Yan Zhou, Fang Zhou Prevalence of self-reported depression and anxiety among pediatric medical staff members during the COVID-19 outbreak in Guiyang, China. Psychiatry Research. Volume 288, June 2020, 113005. https://doi.org/10.1016/j.psychres.2020.113005.

